# Geographical variation and mediation analysis of incidence in stroke: Insights from CHARLS cohort study

**DOI:** 10.1101/2025.04.21.25326174

**Authors:** Yuanyuan Anna Wang, Jiaying Liu, Huazheng Liang

## Abstract

**Introduction:** Stroke is leading cause of mortality in China and it has regional disparities in the incidence among Chinse populations. The study aimed to investigate whether and how much of the association between regional disparities and stroke is explained by an mediator.

**Methods:** We applied Cox proportional hazard regression model and mediation analysis to analyse results from the follow-up of of the CHARLS database.

**Results:** Over the median follow-up of 9.8 years in CHARLS, 5.1 (%) of CHARLS participants developed a stroke. We found that North China had the highest incidence of stroke, followed by Central, Northeast, Northwest, Eastern, and Southwest China. South China had the lowest incidence of stroke, and was the reference group. Rural residents who were older and smokers had a higher risk of stroke. Subgroup analysis revealed that the incidence of stroke in Northeast China was significantly higher among female urban residents with lower education levels, diabetes, and hypertension. The mediation analysis showed that hypertension had a significant mediation effect in Northeast, North, and Northwest China. Dyslipidemia had similar mediating effect in the Northwest and North China, but not in other regions.

**Conclusion:** Geographical variation is associated with a different incidence of stroke. This association is partially mediated by hypertension in Northern, Northeastern, and Northwestern China, and by dyslipidemia in Northwestern and Northern China.

## Introduction

Stroke is the second leading cause of death globally, with approximately 6 million deaths worldwide in 2019^1^. From 1990 to 2019, age-adjusted prevalence of stroke has remained stable in high-income countries, but is increasing in low-income and middle-income countries^2^ There is a considerable degree of geographic variation in the incidence, prevalence, mortality, and DALYs associated with stroke^1, 2^.

In China, stroke has been the leading cause of death^3, 4^, and the prevalence of stroke has been increasing, by 2.2% per year from 2013-2019^4^. Middle-aged and elderly people have a higher prevalence of stroke than other age groups^5^. Since 2011, the Chinese government has initiated a stroke prevention and control project entitled the China Stroke Prevention Project Committee Stroke Program to provide comprehensive interventions for people older than 40 years^6, 7^. Rural regions had a significantly higher prevalence than the urban counterpart^4, 5^. A significant regional difference was found in the prevalence of stroke by 2017^4^.

Many factors are well known to influence the stroke, such as age, gender, hypertension, diabetes and dyslipidemia, etc ^1, 8^. However, as China has a large area and many ethnic groups, there is a significant difference in lifestyle between regions. Geographic differences will change the risk of developing dyslipidemia, diabetes, or hypertension^5, 7^. High-salt, high-carb, high-fat diet in northern China may lead to chronic diseases, such as hypertension, diabetes and hyperlipidemia, which can lead to stroke^4, 9^.

Although previous study conducted a large-scale epidemiological analysis estimating stroke burden in China in 2017^4, 7^, the paper itself does not focus on mediation analysis, such methods could be applied to its data to investigate mechanisms behind regional disparities in stroke incidence. It is more important to explore the pathways through which regional disparities influence an incidence of stroke. Thus, the study aims to disentangle direct and indirect effects, helping researchers understand whether and how much of the association between regional disparities and stroke is explained by an mediator. This is extremely important for regional governments to take target health promotion strategies and improvement for public people inflicted by these mediators.

## 2. Method

### 2.1 Study Population

Data were extracted from the China Health and Retirement Longitudinal Study (CHARLS). CHARLS is an ongoing national-scale and household-based survey of residents aged 45 years or above, launched in 2011 with follow-ups every 1-2 years. By 2020, it covered 19,395 residents across 450 villages/communities in 28 provinces^1^. Data from 2011, 2013, 2015, 2018 and 2020 waves of CHARLS were analysed in this study.

Inclusion criteria included: aged 45 or above; detailed demographic information including age, gender, residence, etc. The following 5 exclusion criteria were applied to screen the participants of CHARLS: 1) missing information on health status and functioning (n = 501), 2) missing information on PSU (Primary Sampling Unit) (n=7), 3) lost data during the follow-up (n = 12,123), 4) age < 45 years (n = 261), and 5) missing information on location, stroke, and covariates (n = 1,672). Finally, 11,625 participants from CHARLS were enrolled in this study (**Fig. S1**). Written informed consent was obtained from all participants. CHARLS was approved by the Ethical Review Committee of Peking University and conducted by the National School for Development (China Centre for Economic Research) at Peking University.

### 2.2 Definitions of Variable

The identification of location is based on the PSU dataset published by CHARLS, which in 2011, 2013, and 2014 made public individual city information, including the names of provinces and prefecture-level cities. Although unpublished after 2015, PSU data remains valid as survey areas are fixed. Based on this, China is divided into seven regions (Northeast, Northwest, West, Central, East, Southwest, and South China) for final location classification in this study.

The method of identification of stroke in this study was self-reported based on physician diagnosis, which has been proven to be consistent with medical records^10^. Participants were asked: “Have you ever been diagnosed with stroke by a doctor?”. The respondents have reported stroke if they answered “yes” to the question.

Hypertension, diabetes, and dyslipidemia were included as potential mediators between location and stroke. Comorbidities were assessed using: “Have you been diagnosed with hypertension / dyslipidemia (elevation of low-density lipoprotein, triglycerides (TGs), and total cholesterol, or a low high density lipoprotein level) / diabetes or high blood sugar by a doctor?”. Demographic and lifestyle adjustments included age (≥45 years), gender (male, female), residence (rural, urban), education (illiterate, literate and primary, middle school education or above), and smoking status (current, former and never smoker). Standardized questionnaires were used for data collection.

### 2.4 Statistical analysis

Baseline characteristics of the participants were presented as mean and standard deviation (SD) for continuous variables and percentage (%) for categorical variables, with comparisons made using t-test (for continuous variables) and Chi square test (categorical variables), respectively.

The Cox proportional hazard regression model was used to estimate the hazard ratio (HR) and 95% confidence interval of stroke. We used four stepwise adjustments models to determine the link between location and stroke: Model 1 was an unadjusted rough model, which only included location and stroke; Models 2, 3, and 4 adjusted for demographic characteristics, lifestyle, and comorbidities, respectively, based on the previous model.

Stratified analyses were performed across subgroups to assess potential variations, with adjustments for all covariates and mediators. To explore the underlying mechanism, mediation analysis tested whether hypertension, diabetes, and dyslipidemia mediated the association between location (five areas) and stroke incidence. The difference method was used to calculate the mediation proportion by the mediators (hypertension, diabetes and dyslipidemia, respectively) for the association between location and stroke^11^.

All analyses were performed using Stata 15. All P values were two-sided, and statistical significance was defined as P< 0.05.

## 3. Results

### 3.1 Population characteristics

Demographic characteristics of participants included in the present study from CHARLS were presented in **Table 1**. The average age of these 11,625 participants was 57.89 years. In the present study, 1.73% of participants had stroke, 22.48% had hypertension, 5.03% had diabetes and 8.92% had dyslipidemia. North China had the highest incidence of stroke (2.84%). The incidence of stroke was lower in Eastern China (1.32%), Southwest China (1.25%) and Southern China (1.24%), with the lowest in South China at 1.24%. Therefore, we set South China as the reference group to study the differences in other regions. All demographic factors and comorbidities were significantly associated with stroke: Spearman correlation coefficients were all significant at the 95% level (**Fig. S2**).

**Table 1.** Baseline characteristics of participants from CHARLS^a^.

| Characteristics | Total (%) | Northeast China (%) | North China (%) | Northwest China (%) | Central China (%) | East China (%) | Southwest China (%) | South China (%) | P values |
| --- | --- | --- | --- | --- | --- | --- | --- | --- | --- |
| Age (years); Mean (SD<sup>c</sup>) | 57.89 (8.63) | 57.31 (8.24) | 56.71 (8.14) | 56.59 (8.11) | 57.78 (8.55) | 58.43 (8.93) | 58.49 (8.62) | 58.52 (8.99) |  |
| (years) |  |  |  |  |  |  |  |  |  |
| Gender |  |  |  |  |  |  |  |  | 0.960 |
| Male | 5442 (46.82) | 325 (45.45) | 728 (47.93) | 377 (46.89) | 1165 (46.88) | 1279 (46.90) | 970 (46.66) | 598 (46.25) |  |
| Female | 6180 (53.18) | 390 (54.55) | 791 (52.07) | 427 (53.11) | 1320 (53.12) | 1448 (53.10) | 1109 (53.34) | 695 (53.75) |  |
| Residence |  |  |  |  |  |  |  |  | 0.000 |
| Rural | 7608 (65.45) | 403 (56.36) | 991(65.24) | 618 (76.87) | 1556 (62.59) | 1754 (64.32) | 1462 (70.25) | 824 (63.73) |  |
| Urban | 4017 (34.55) | 312 (43.64) | 528 (34.76) | 186 (23.13) | 930 (37.41) | 973 (35.68) | 619 (29.75) | 469 (36.27) |  |
| Education level |  |  |  |  |  |  |  |  | 0.000 |
| Illiterate | 3108 (26.76) | 86 (12.03) | 233 (15.36) | 259 (32.25) | 595 (23.93) | 911 (33.46) | 672 (32.35) | 352 (27.22) |  |
| Literate and primary education | 4687 (40.36) | 313 (43.78) | 563 (37.11) | 269 (33.50) | 978 (39.34) | 966 (35.48) | 1022 (49.21) | 576 (44.55) |  |
| Middle school education or above | 3819 (32.88) | 316 (44.20) | 721 (47.53) | 275 (34.25) | 913 (36.73) | 846 (31.07) | 383 (18.44) | 365 (28.23) |  |
| Smoking status |  |  |  |  |  |  |  |  | 0.000 |
| Never | 7119 (61.24) | 395 (55.24) | 881 (58.00) | 479 (59.58) | 1542 (62.03) | 1680 (61.63) | 1317 (63.29) | 825 (63.81) |  |
| Current | 3652 (31.42) | 266 (37.20) | 508 (33.44) | 271 (33.71) | 743 (29.89) | 808 (29.64) | 654 (30.70) | 402 (31.09) |  |
| Former | 853 (7.34) | 54 (7.55) | 130 (8.56) | 54 (6.72) | 201 (8.09) | 238 (8.73) | 110 (5.29) | 66 (5.10) |  |
| stroke |  |  |  |  |  |  |  |  |  |
| Yes | 201 (1.73) | 12 (1.69) | 43 (2.84) | 11 (1.37) | 57 (2.30) | 36 (1.32) | 26 (1.25) | 16 (1.24) |  |
| No | 11399<br>(98.27) | 700<br>(98.31) | 1473<br>(97.16) | 794<br>(98.63) | 2426<br>(97.70) | 2686<br>(98.68) | 2053<br>(98.75) | 1286<br>(98.76) |  |
| Hypertension |  |  |  |  |  |  |  |  | 0.000 |
| Yes | 2598<br>(22.48) | 186<br>(26.09) | 464<br>(30.85) | 192<br>(23.94) | 499<br>(20.13) | 730<br>(26.89) | 353<br>(17.14) | 174<br>(13.52) |  |
| No | 8961<br>(77.52) | 527<br>(73.91) | 1504<br>(69.15) | 610<br>(76.06) | 2037<br>(80.07) | 1985<br>(73.11) | 1706<br>(82.86) | 1113<br>(86.48) |  |
| Diabetes |  |  |  |  |  |  |  |  | 0.000 |
| Yes | 580<br>(5.03)1 | 47<br>(6.60) | 98<br>(6.50) | 30<br>(3.77) | 121<br>(4.88) | 184<br>(6.82) | 66<br>(3.19) | 34<br>(2.66) |  |
| No | 10953<br>(94.97) | 665<br>(93.40) | 1409<br>(93.50) | 765<br>(96.23) | 2357<br>(95.12) | 2512<br>(93.18) | 2001<br>(96.81) | 1244<br>(97.34) |  |
| Dyslipidemia |  |  |  |  |  |  |  |  | 0.000 |
| Yes | 1017<br>(8.92) | 91<br>(12.83) | 215<br>(14.49) | 71<br>(9.20) | 230<br>(9.12) | 294<br>(11.07) | 61<br>(2.97) | 60<br>(4.75) |  |
| No | 10390<br>(91.08) | 618<br>(87.17) | 1269<br>(85.51) | 701<br>(90.80) | 2243<br>(90.88) | 2363<br>(88.93) | 1994<br>(97.03) | 1202<br>(95.25) |  |

<sup>a</sup>:Values are numbers (percentages) unless stated otherwise;
<sup>b</sup>: CHARLS: The China Health and Retirement Longitudinal Study;
<sup>c</sup>:SD: standard deviation.

### 3.2 Regression Analysis

Over the median follow-up of 9.8 year in CHARLS, 5.1 (%) of CHARLS participants developed stroke. **Table 2** illustrated the cox proportional hazards regression effect of location on the incidence of stroke. Our findings suggest that the correlation between stroke and location was significant in all four models. It can be seen that the inclusion of demographic factors and lifestyle covariates decreased the hazard ratio in some areas (Northeast China: HR from 3.68 to 3.66, North China: HR from 3.24 to 3.22, Central China: HR from 1.74 to 1.73). After including all covariates, the hazard ratios (HR) for stroke incidence in different areas were still significant.

**Table 2.**
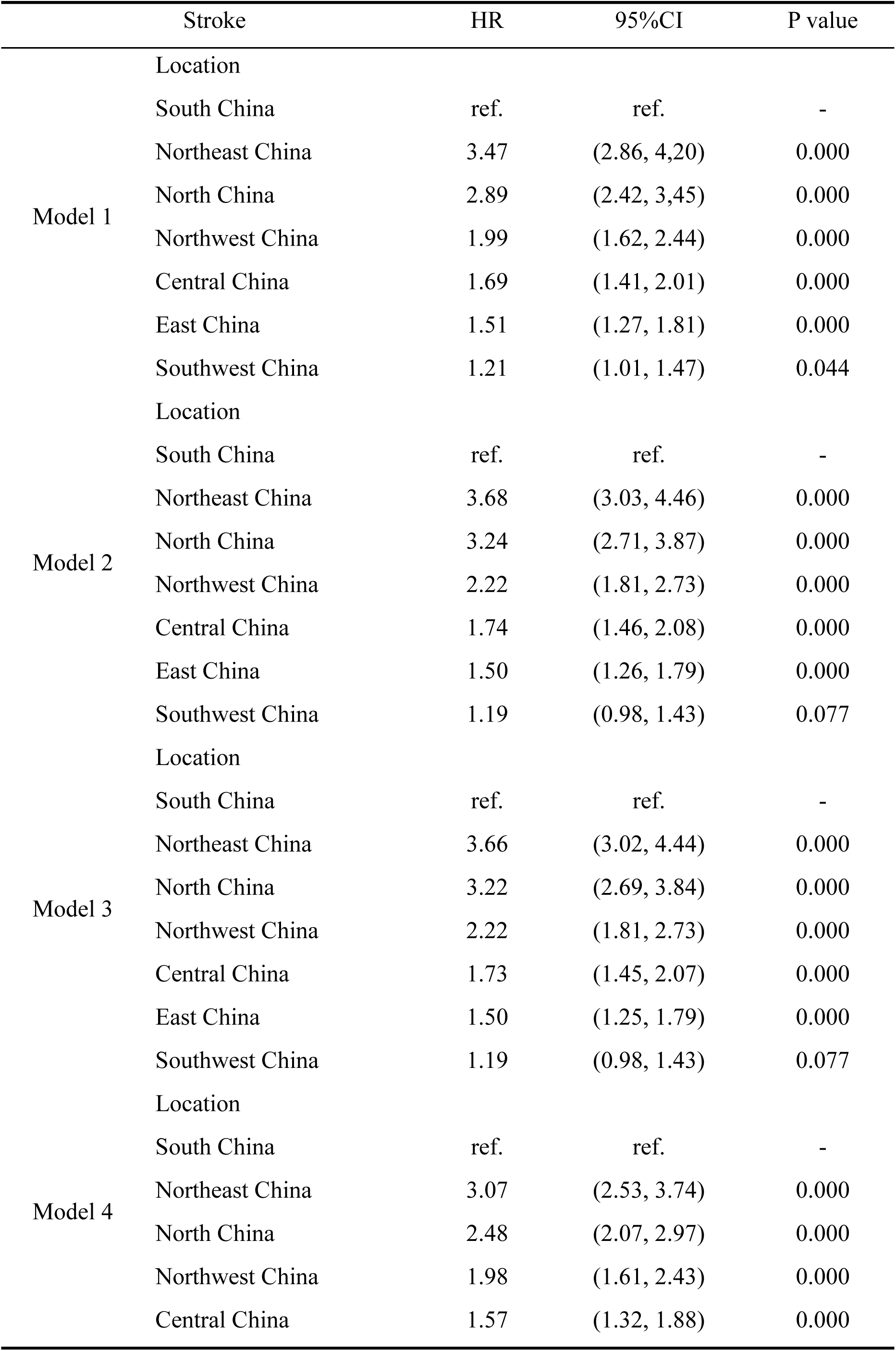
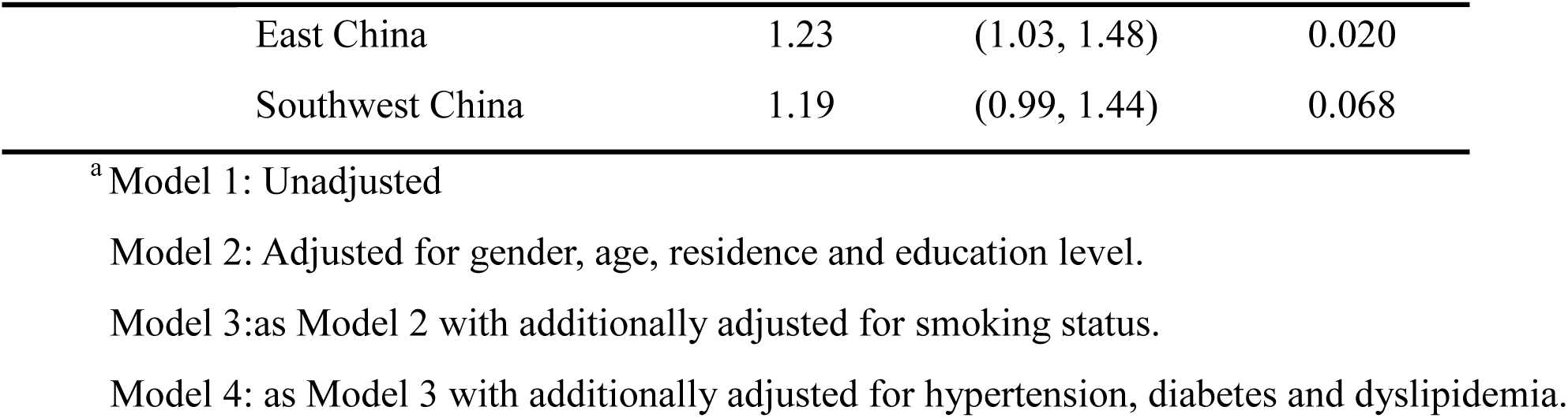
Association between location and the risk of stroke in the CHARLS.

|  | Stroke | HR | 95%CI | P value |
| --- | --- | --- | --- | --- |
| Model 1 | Location |  |  |  |
|  | South China | ref. | ref. | - |
|  | Northeast China | 3.47 | (2.86, 4.20) | 0.000 |
|  | North China | 2.89 | (2.42, 3.45) | 0.000 |
|  | Northwest China | 1.99 | (1.62, 2.44) | 0.000 |
|  | Central China | 1.69 | (1.41, 2.01) | 0.000 |
|  | East China | 1.51 | (1.27, 1.81) | 0.000 |
|  | Southwest China | 1.21 | (1.01, 1.47) | 0.044 |
| Model 2 | Location |  |  |  |
|  | South China | ref. | ref. | - |
|  | Northeast China | 3.68 | (3.03, 4.46) | 0.000 |
|  | North China | 3.24 | (2.71, 3.87) | 0.000 |
|  | Northwest China | 2.22 | (1.81, 2.73) | 0.000 |
|  | Central China | 1.74 | (1.46, 2.08) | 0.000 |
|  | East China | 1.50 | (1.26, 1.79) | 0.000 |
|  | Southwest China | 1.19 | (0.98, 1.43) | 0.077 |
| Model 3 | Location |  |  |  |
|  | South China | ref. | ref. | - |
|  | Northeast China | 3.66 | (3.02, 4.44) | 0.000 |
|  | North China | 3.22 | (2.69, 3.84) | 0.000 |
|  | Northwest China | 2.22 | (1.81, 2.73) | 0.000 |
|  | Central China | 1.73 | (1.45, 2.07) | 0.000 |
|  | East China | 1.50 | (1.25, 1.79) | 0.000 |
|  | Southwest China | 1.19 | (0.98, 1.43) | 0.077 |
| Model 4 | Location |  |  |  |
|  | South China | ref. | ref. | - |
|  | Northeast China | 3.07 | (2.53, 3.74) | 0.000 |
|  | North China | 2.48 | (2.07, 2.97) | 0.000 |
|  | Northwest China | 1.98 | (1.61, 2.43) | 0.000 |
|  | Central China | 1.57 | (1.32, 1.88) | 0.000 |

|  |  |  |  |
| --- | --- | --- | --- |
| East China | 1.23 | (1.03, 1.48) | 0.020 |
| Southwest China | 1.19 | (0.99, 1.44) | 0.068 |

<sup>a</sup> Model 1: Unadjusted
Model 2: Adjusted for gender, age, residence and education level.
Model 3: as Model 2 with additionally adjusted for smoking status.
Model 4: as Model 3 with additionally adjusted for hypertension, diabetes and dyslipidemia.

Stratified analyses (Fig. 1) revealed significant variations in stroke incidence across regions and demographic groups. In Northeast China, stroke incidence was higher in urban areas, women, individuals with lower education levels, and those with diabetes or dyslipidemia. North China exhibited a similar pattern, except stroke incidence was higher in rural than urban areas. In Eastern China, older adults had elevated stroke rates, while Southwest China showed significantly higher incidence among rural residents.

### 3.3 Mediation analysis

As seen in **Table 3**, when hypertension was used as a mediating variable, its mediating effect was prevalent in Northern region (**Fig. S3A**), with the mediator proportion of 1.55% (95% CI: 0.18, 2.56), 9.46% (95% CI: 9.41, 9.49) and 8.57% (95% CI: 8.33, 8.84) in Northeast, North, and Northwest China, respectively. In the eastern region, the effect of hypertension on the incidence of stroke was a masking effect (**Fig. S3B**). Secondly, in Northwest and Eastern China, diabetes acted as a significant masking effect on stroke (**Fig. S3B**). Finally, the proportion of mediators of dyslipidemia was higher in the northwest and northern regions (**Fig. S3A**), with mediator proportions of 2.13% (95%CI: 0.01, 2.69), and 5.64% (95%CI: 5.20, 5.88), respectively. In the eastern region, the effect of dyslipidemia on the incidence of stroke presented as a masking effect (**Fig. S3B**). Thus, blood pressure control is a key factor in stroke prevention, and the effect is more pronounced in the North China. In addition, as hypertension and dyslipidemia both mediated the effect of northern region on the incidence of stroke, we performed a parallel mediation test **(Table S2)**.

**Table 3.** Direct, indirect effects and mediation proportions between location and stroke mediated by hypertension, diabetes and dyslipidemia.

|  | Direct effect<sup>a</sup> |  | Indirect effect<sup>a</sup> |  | Proportion Mediated |
| --- | --- | --- | --- | --- | --- |
| | $\beta^b$ | (95%CI<sup>c</sup>) | $\beta^b$ | (95%CI<sup>c</sup>) | % (95%CI<sup>c</sup>) |
| Hypertension |  |  |  |  |  |
| South China | ref. | ref. | ref. | ref. | - |
| Northeast China | 0.064 | (0.055, 0.076) | 0.001 | (0.000, 0.002) | 1.55(0.18, 2.56) |
| North China | 0.060 | (0.051, 0.071) | 0.006 | (0.005, 0.007) | 9.46(9.41, 9.49) |
| Northwest China | 0.013 | (0.005, 0.022) | 0.001 | (0.000, 0.002) | 8.57(8.33, 8.84) |
| Central China | -0.001 | (-0.001, 0.007) | -0.003 | (-0.004, 0.002) | - |
| East China | -0.037 | (-0.045, -0.031) | 0.005 | (0.004, 0.006) | -14.20(-22.49, -8.98) |
| Diabetes |  |  |  |  |  |
| South China | ref. | ref. | ref. | ref. | - |
| Northeast China | 0.064 | (0.054, 0.075) | -0.000 | (-0.000,0.000) | - |
| North China | 0.060 | (0.050, 0.069) | 0.000 | (-0.000, 0.000) | - |
| Northwest China | 0.013 | (0.004, 0.022) | -0.000 | (-0.000, -0.000) | -1.55(-3.16, 0.00) |
| Central China | -0.001 | (-0.009, 0.008) | -0.000 | (-0.000, 0.000) | - |
| East China | -0.037 | (-0.044, -0.030) | 0.001 | (0.000, 0.001) | -2.22(-3.16, -1.23) |
| Dyslipidemia |  |  |  |  |  |
| South China | ref. | ref. | ref. | ref. | - |
| Northeast China | 0.064 | (0.055, 0.076) | 0.001 | (0.001, 0.002) | 2.13(0.01, 2.69) |
| North China | 0.060 | (0.051, 0.070) | 0.004 | (0.003, 0.004) | 5.64(5.20, 5.88) |
| Northwest China | 0.013 | (0.005, 0.022) | 0.000 | (-0.000, 0.009) | - |
| Central China | -0.001 | (-0.008, 0.007) | 0.001 | (0.000, 0.001) | - |
| East China | -0.037 | (-0.044, -0.029) | 0.001 | (0.000, 0.002) | -2.29(-6.87, -0.76) |

<sup>a</sup> Adjusted models include the following covariates: gender, age, residence, education level, smoking status, hypertension, diabetes and dyslipidemia.
<sup>b</sup> $\beta$ : bootstrap mediation effect test coefficients
<sup>c</sup> CI: bias-corrected confidence interval

### 3.4 Sensitivity Analyses

Sensitivity analyses showed that the main results were robust (**Table S1**). Firstly, similar results were obtained when Logit was used to replace the cox regression model. Secondly, the inclusion of respondents younger than 45 years old yielded results that were broadly similar to those of the main analysis, suggesting that our main findings are not biased by baseline characteristics of the subjects. Finally, the missing data were interpolated multiple times to test the effect of missing variables, and the results were generally consistent with the main analysis.

## Discussion

The study found that the incidence of stroke was highest in northern China and lowest in southern China. Rural residents who were older and smokers with lower education levels, diabetes, and hypertension had a higher risk of stroke. Hypertension took a significant mediation effect in Northeast, North China, and Northwest China. Dyslipidemia took such an effect in the Northwest and North China.

North, Central and Northeast China showed the higher incidence of stroke as reported in this study, which is similar with other studies^4, 12^. In an over 1.8 million-admission study^13^, stroke in North China were the youngest in average and more likely to have severe comorbidities. It was assumed that differences in lifestyle^4^, environment^14^, economy level and medical development^15^ may account for such variation between regions.

The increasing incidence of stroke in China from 1990 to 2016 to 2021 raises a strong need to prevent stroke by taking appropriate measures^7, 16^. Hypertension was found to be a significant mediator of stroke^17^. But our study is the first one to investigate the mediation effect of key factors of stroke and found that blood pressure (BP) control was more important in Northeast, North and Northwest China. In addition, dyslipidemia results in deposition of lipids in blood vessels and subsequent artheriosclerosis, which may eventually thrombosis. So controlling lipids is also very important in controlling stroke^18^.

There are also several limitations of this study. First, due to limitations in data availability, we could only include self-reported stroke by participants. This could lead to an underestimation of the incidence of stroke^19^. Second, there is a partial loss of data for some participants during the follow-up. It could result in selection bias^20^. Finally, since the CHARLS data is only updated to 2020 and data after the COVID-2019 is not currently available, the impact on the habits and physical status of the respondents regarding the epidemic is unknown.

### Conclusions

This large-scale Chinese cohort study revealed a north-to-south geographical gradient in stroke incidence, with higher rates in northern regions. Hypertension and dyslipidemia mediated this regional disparity, particularly in the north. Older rural residents and smokers faced elevated stroke risks. The finding highlights the importance of management of hypertension and dyslipidemia among people live in northern, northeast and northwest China, which may tackle the incidence of stroke.

## Data Availability

Data are available from the CHARLS database website and also available from the corresponding author upon request.

https://charls.charlsdata.com

## Acknowledgments

The authors appreciate the support of participants and colleagues in the same institute.

## Authors’ contributions

YW and HL conceived and designed the study. YW and YL performed data analysis and wrote the draft. YL, HL, and YW contributed to data analysis. YL reviewed the statistical methods and results. YW and HL critically revised the manuscript. All authors read and approved the final manuscript.

## Consent for publication

Not applicable.

---

**SUPPLEMENTARY FIGURE S1.**
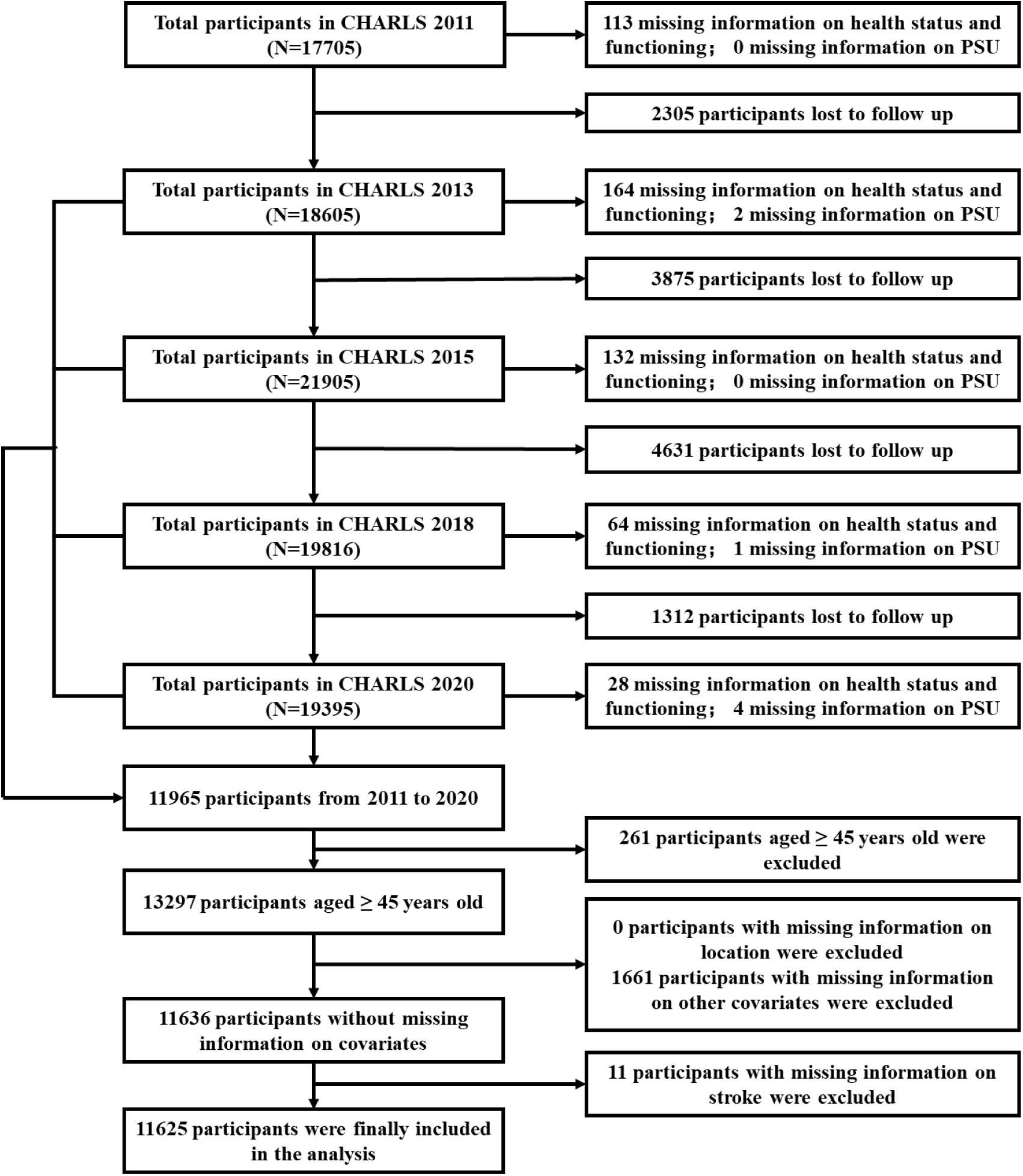
Flowchart of the study sample.

**SUPPLEMENTARY FIGURE S2.**
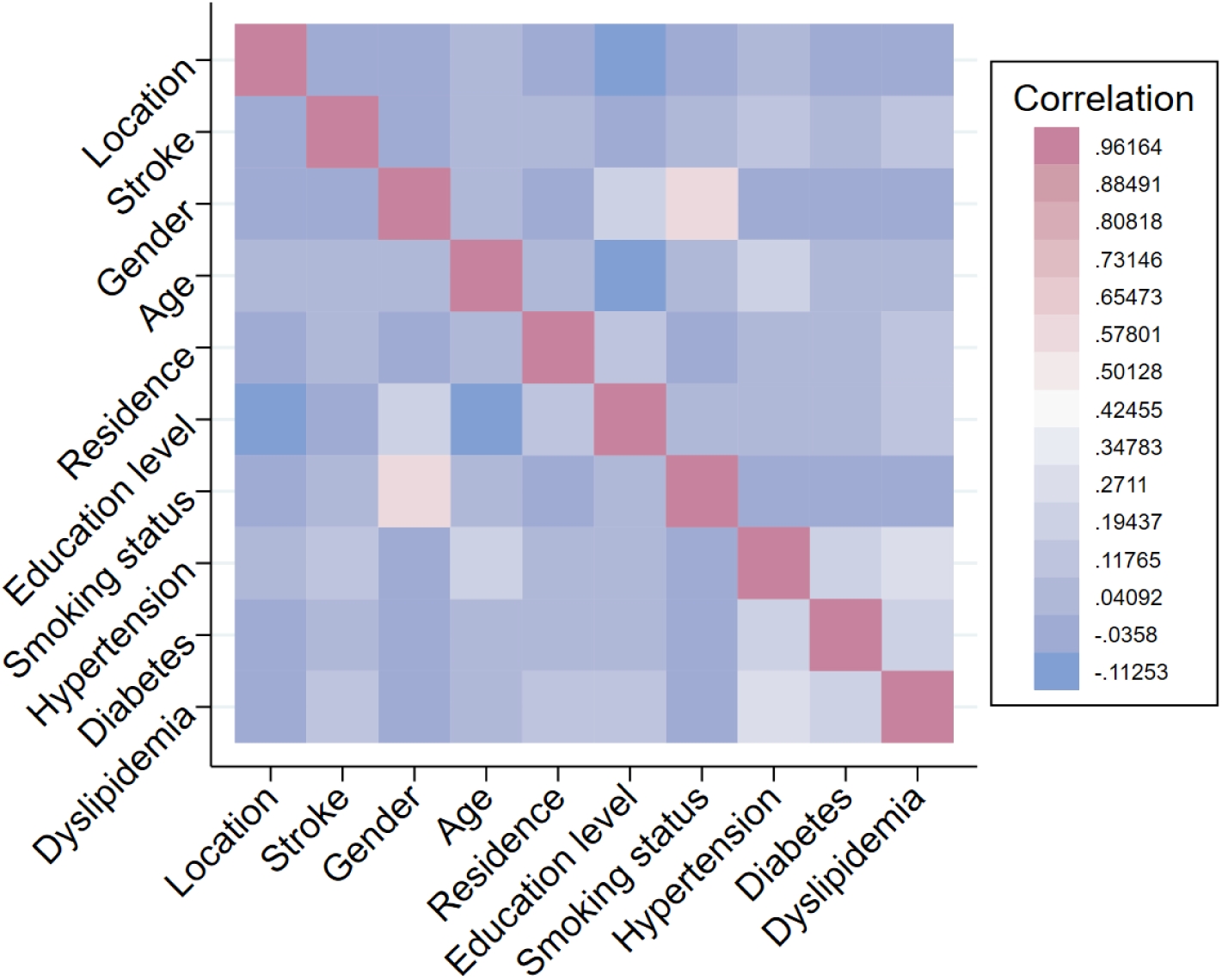
Correlation coefficient of explanatory variables.

**SUPPLEMENTARY FIGURE S3.**
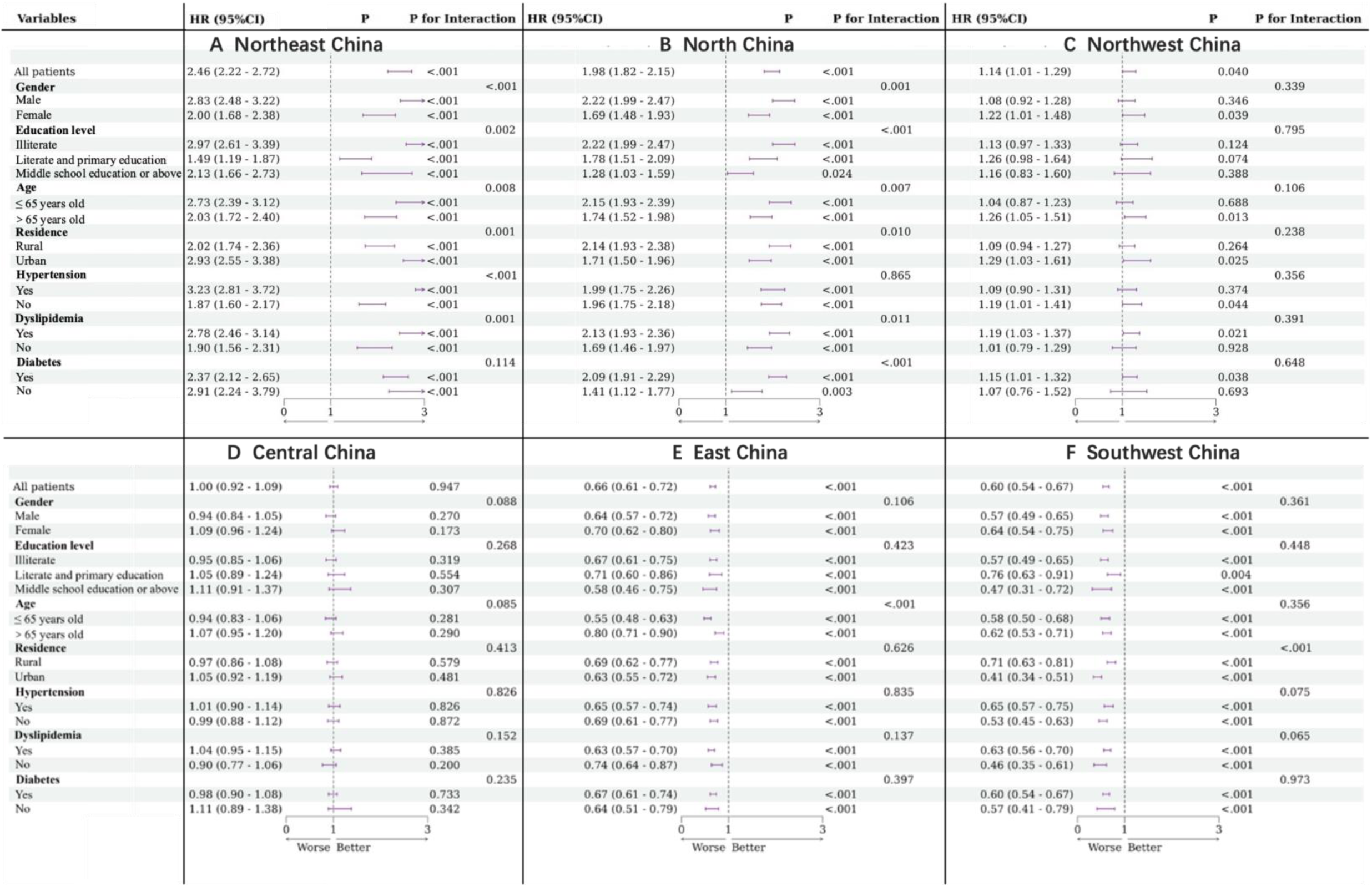
Adjusted hazard ratios (95% confidence intervals) for incidence of stroke stratified by gender, education level, age, residence, hypertension, dyslipidemia, diabetes

**SUPPLEMENTARY FIGURE S4.**
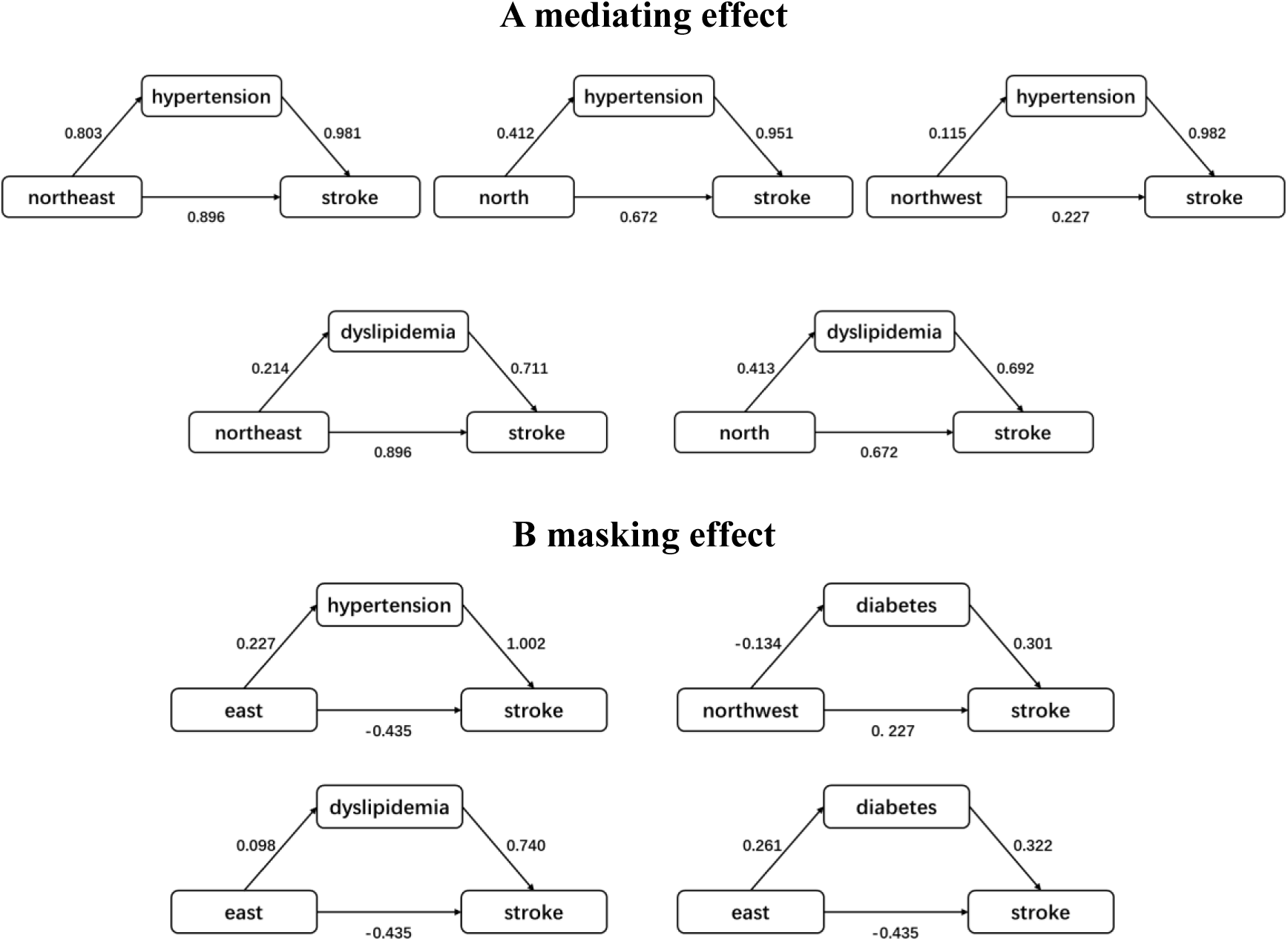
^a^ Schematic representation of the mediating effect and masking effect of location on stroke using hypertension, diabetes, and dyslipidemia as mediating variables. ^a^The data in the figure refers to the mediation effect two-step coefficients.

**SUPPLEMENTARY TABLE S1.** Associations of diabetes with incident chronic lung diseases: sensitivity analyses*.

| Analyses | Estimated using the Logit model | | | Among individuals $\leq 45$ years | | | multiple imputation | | |
| --- | --- | --- | --- | --- | --- | --- | --- | --- | --- |
|  | OR | 95%CI | P value | HR | 95%CI | P value | HR | 95%CI | P value |
| South China | ref. | ref. | ref. | ref. | ref. | ref. | ref. | ref. | ref. |
| Northeast China | 4.09 | (3.41, 4.90) | 0.000 | 3.11 | (2.54, 3.80) | 0.000 | 3.10 | (2.55, 3.77) | 0.000 |
| North China | 3.12 | (2.64, 3.69) | 0.000 | 2.50 | (2.07, 3.01) | 0.000 | 2.53 | (2.11, 3.02) | 0.000 |
| Northwest China | 2.16 | (1.79, 2.63) | 0.000 | 1.99 | (1.60, 2.47) | 0.000 | 1.98 | (1.62, 2.43) | 0.000 |
| Central China | 1.74 | (1.47, 2.05) | 0.000 | 1.58 | (1.31, 1.89) | 0.000 | 1.60 | (1.34, 1.91) | 0.000 |
| East China | 1.25 | (1.05, 1.48) | 0.009 | 1.23 | (1.03, 1.49) | 0.026 | 1.27 | (1.07, 1.52) | 0.007 |
| Southwest China | 1.17 | (0.98, 1.40) | 0.084 | 1.21 | (0.99, 1.47) | 0.060 | 1.23 | (1.02, 1.48) | 0.031 |

\*Adjusted models include the following covariates: gender, age, residence, education level, smoking status, hypertension, diabetes and dyslipidemia.

**SUPPLEMENTARY TABLE S2.** Parallel mediation effects between North China and stroke mediated by hypertension and dyslipidemia.

|  | Direct effect<sup>a</sup> |  | Indirect effect<sup>a</sup> |  | Proportion Mediated (%) (CI95%) |
| --- | --- | --- | --- | --- | --- |
|  | OR | (CI<sup>b</sup> 95%) | OR | (CI<sup>b</sup> 95%) |  |
| Hypertension | - | - | 0.005 | (0.005, 0.006) | 10.47(10.33, 10.57) |
| Dyslipidemia | - | - | 0.004 | (0.003, 0.004) | 7.54(7.31, 7.71) |
| Total | 0.041 | (0.036, 0.046) | 0.009 | (0.008, 0.010) | 18.01(17.69,18.42) |

<sup>a</sup> Adjusted models include the following covariates: gender, age, residence education level, smoking status, hypertension and dyslipidemia.
<sup>b</sup> CI: bias-corrected confidence interval

## Footnotes

1 Further details about the study design and implementation procedures of CHARLS are available at the official website of CHARLS (http://charls.pku.edu.cn/en).

